# Getting to zero quickly in the 2019-nCov epidemic with vaccines or rapid testing

**DOI:** 10.1101/2020.02.03.20020271

**Authors:** Gerardo Chowell, Ranu Dhillon, Devabhaktuni Srikrishna

## Abstract

Any plan for stopping the ongoing 2019-nCov epidemic must be based on a quantitative understanding of the proportion of the at-risk population that needs to be protected by effective control measures in order for transmission to decline sufficiently and quickly enough for the epidemic to end. Using an SEIR-type transmission model, we contrasted two alternate strategies by modeling the proportion of the population that needs to be protected from infection by one-time vaccination (assuming 100% effectiveness) or by testing with isolation and treatment of individuals within six, 24, or 48 hours of symptom onset. If R is currently 2.2, vaccination at the herd immunity coverage of 55% would drive R just below 1, but transmission could persist for years. Over 80% of coverage is required to end the epidemic in 6 months with population-wide vaccination. The epidemic could be ended in just under a year if testing with isolation and treatment reached 80% of symptomatically infected patients within 24 hours of symptom onset (assuming 10% asymptomatic transmission). The epidemic could be ended in six months if testing with isolation and treatment reached 90% of symptomatic patients. If 90% of symptomatic patients could be tested within six hours of symptoms appearing, the epidemic could be ended in under four months.

---

Any plan for stopping the ongoing 2019-nCov epidemic must be based on a quantitative understanding of the proportion of the at-risk population that needs to be protected by effective control measures in order for transmission to decline sufficiently and quickly enough for the epidemic to end.

Ending the epidemic requires reducing the basic reproduction number (R) – the average number of people infected by each 2019-nCov-infected patient – to below 1, at which point transmission contracts and eventually burns out. However, if R is only brought marginally below 1, transmission could still linger for years. Enough infections could occur during this time that, especially given high mobility of people and density of urban areas, new hotspots could get seeded and transmission could resurge above 1. Ending the epidemic, therefore, may require not just getting R below 1, but driving it down more aggressively in order to reach zero cases quickly.

In January, the mean R was estimated at 2.2 (3.9, upper bound of 95% confidence interval), with each infection on average resulting in two or more infections [1]. Although cases of asymptomatic transmission have been documented [2,3], the World Health Organization advises that transmission is still primarily driven by symptomatic patients [4].

In order to bring R below 1, we contrasted two alternate strategies by modeling the proportion of the population that needs to be protected from infection by one-time vaccination (assuming 100% effectiveness) or by testing with isolation and treatment of individuals within six, 24, or 48 hours of symptom onset. We employed an SEIR-type transmission model [5] where the mean incubation period is 5 days, mean infectious period is 7 days, and the initial prevalence of the novel coronavirus (2019-nCov) infections is at 0.1% in a population of 10 million, roughly the population of Wuhan. In light of the unknowns with asymptomatic transmission, a fixed percentage of the transmission is modeled prior to symptom onset.

If the current R is 2.2 or 3.9, 55% or 74% of the at-risk population, respectively, would need to be vaccinated in order to bring R below 1. If R is 2.2 and 10% of transmission occurs asymptomatically, approximately 60%, 68%, or 82% of symptomatic individuals would need to be tested and isolated for treatment within six, 24, or 48 hours, respectively, of symptom onset in order to bring R below 1. If asymptomatic transmission accounted for 20% of overall transmission, coverage would need to reach 65%, 74%, and 87% of the symptomatic individuals, respectively. If R is 3.9 and 10% of transmission occurs asymptomatically, testing with isolation and treatment would need to reach 83% or 93% of symptomatic individuals if done within six or 24 hours of symptom onset, respectively. Given that the denominator for population-wide vaccination (∼10 million) is much greater than the proportion of that population with concerning symptoms, far fewer people would need to be reached with testing and isolation for treatment than for vaccinating to achieve herd immunity.

We then modeled the number of days it would take to get to zero with different rates of coverage with vaccination or testing with isolation and treatment of symptomatically infected patients (Figure 1). If R is currently 2.2, for example, vaccination at the herd immunity coverage of 55% would drive R just below 1, but transmission could persist for years. Over 80% of coverage is required to end the epidemic in 6 months with population-wide vaccination. The epidemic could be ended in just under a year if testing with isolation and treatment reached 80% of symptomatically infected patients within 24 hours of symptom onset (assuming 10% asymptomatic transmission). The epidemic could be ended in six months if testing with isolation and treatment reached 90% of symptomatic patients. If 90% of symptomatic patients could be tested within six hours of symptoms appearing, the epidemic could be ended in under four months.

**Figure 1.**
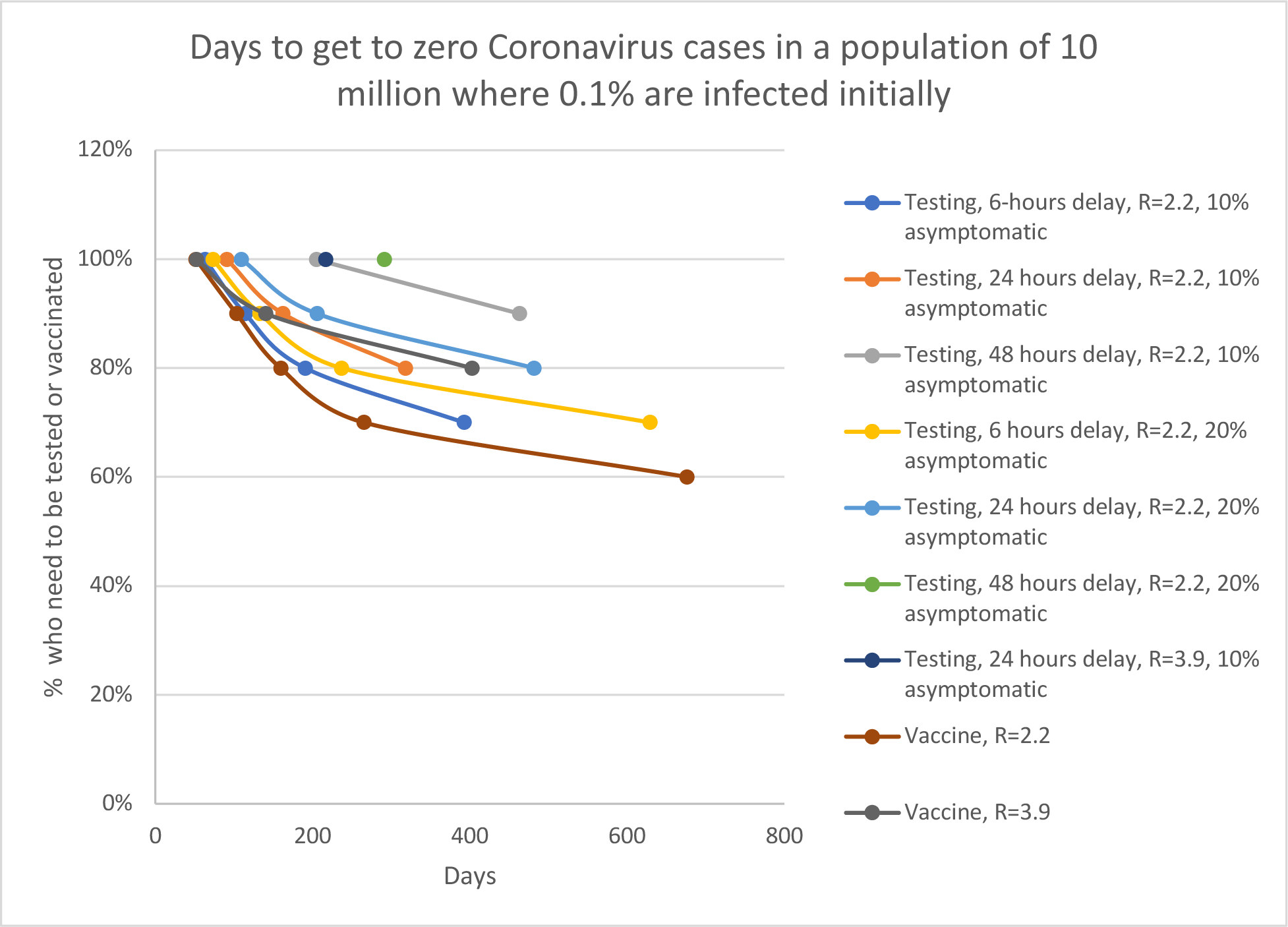
Number of days to get to zero at different rates of vaccine coverage compared to testing followed by treatment/isolation of symptomatic patients.

This epidemic could be ended within months if widely usable diagnostics or vaccines could be developed and used to cover a sufficient proportion of the at-risk population.

## Data Availability

All data is publicly available

### Appendix

#### Basic reproduction number, R_0_

The basic reproduction number is defined as the average number of secondary cases generated by primary infectious individuals during the early transmission phase in a completely susceptible population[1]. This a key metric to gauge the intensity and type of interventions that need to be implemented in order to bring the epidemic under control. For the ongoing epidemic of 2019-nCov, R0 has been estimated at 2.2 (3.9, upper bound of 95% confidence interval).

#### Modeling the effect of testing and treatment/isolation of individuals

We employed an SEIR-type transmission model to evaluate the effects of the percentage of symptomatic individuals that are tested/screened as well as the average time that it takes to bring them into isolation and treatment [2]. The population is classified into 5 epidemiological states: susceptible (S), exposed (E), symptomatic and infectious individuals that are tested (Is), symptomatic infectious individuals that are not tested and remain in the community (In), symptomatic individuals that are effectively isolated and treated (J), and recovered individuals (R). A schematic diagram of the model is given in Figure S1.

Susceptible individuals may be infected by contacting symptomatic and infectious individuals (Is+ In) and partially infectious individuals incubating the disease (E). The force of infection is given by:

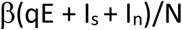

Where β is the transmission rate and q is a parameter ranging from 0 to 1 that models the relative infectiousness of individuals during the incubation period of the disease.

Exposed individuals progress to the symptomatic phase after an average of 5 days whereas the average infectious period for symptomatic individuals is 7 days. The percentage of the symptomatic individuals that undergo testing (Is) ranges from 0 to 100%. The average time from onset of symptoms to isolation and treatment for individuals that undergo testing ranges from 6 to 48 hours.

The reproduction number that includes the effects of testing and isolation and treatment of symptomatic individuals is given by:

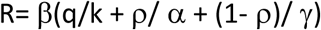

In the absence of testing, the basic reproduction number (R_0_) is simply given by

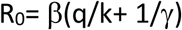

#### Modeling the effect of vaccination

We define the vaccination reproduction number (**R_v_**) for a given effective vaccination coverage level (**v**) as follows:

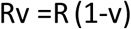

Where v is proportion of the population that is effectively vaccinated.

#### Modeling the time to get to zero

We modeled the average number of days it would take to get to zero cases once the reproduction number declines below the epidemic threshold of 1.0 for various control interventions. For this purpose, we generated stochastic simulations[3] using the compartmental model described in the previous section assuming that the initial prevalence of the novel coronavirus (2019-nCov) infections is at 0.1% in a population of 10 million. To calculate the average number of days to get to zero cases, we simulated a total of 200 stochastic epidemics.

**Table S1.** Parameter definitions and baseline values.

| Parameter | Definition | Baseline value |
| --- | --- | --- |
| $\beta$ | Transmission rate | Calibrated for $R_0=2.2$ or $R_0=3.9$ |
| q | Relative infectiousness of infected individuals incubating the disease | 0.1, 0.2, 0.4 |
| $1/k$ | Incubation period | 5 days |
| $1/\gamma$ | Infectious period | 7 days |
| $1/\alpha$ | Time to isolation/treatment | 6, 24, 48 hours |
| $\rho$ | Proportion of the symptomatic and infectious individuals that undergo testing/screening | 0-100% |
| v | Proportion of the population that is vaccinated | 0-100% |
| N | Population size | 10 million |

**Figure S1.**
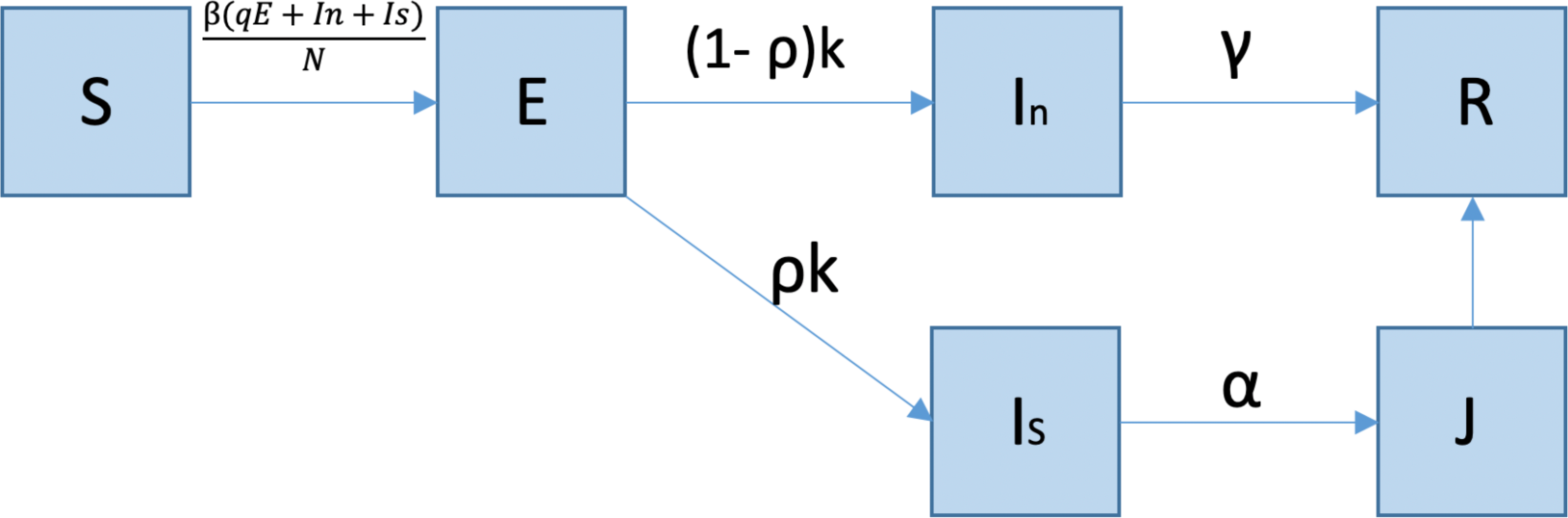
Schematic of the model diagram. The population is classified into 5 epidemiological states: susceptible (S), exposed (E), symptomatic and infectious individuals that are tested (Is), symptomatic infectious individuals that are not tested and remain in the community (In), symptomatic individuals that are effectively isolated and treated (J), and recovered individuals (R). Model parameters are described in Table 1.

